# Urban Air Pollution May Enhance COVID-19 Case-Fatality and Mortality Rates in the United States

**DOI:** 10.1101/2020.05.04.20090746

**Authors:** Donghai Liang, Liuhua Shi, Jingxuan Zhao, Pengfei Liu, Joel Schwartz, Song Gao, Jeremy Sarnat, Yang Liu, Stefanie Ebelt, Noah Scovronick, Howard H Chang

## Abstract

**Background:** The novel human coronavirus disease 2019 (COVID-19) pandemic has claimed more than 240,000 lives worldwide, causing tremendous public health, social, and economic damages. While the risk factors of COVID-19 are still under investigation, environmental factors, such as urban air pollution, may play an important role in increasing population susceptibility to COVID-19 pathogenesis.

**Methods:** We conducted a cross-sectional nationwide study using zero-inflated negative binomial models to estimate the association between long-term (2010-2016) county-level exposures to NO_2_, PM_2.5_ and O_3_ and county-level COVID-19 case-fatality and mortality rates in the US. We used both single and multipollutant models and controlled for spatial trends and a comprehensive set of potential confounders, including state-level test positive rate, county-level healthcare capacity, phase-of-epidemic, population mobility, sociodemographic, socioeconomic status, behavior risk factors, and meteorological factors.

**Results:** 1,027,799 COVID-19 cases and 58,489 deaths were reported in 3,122 US counties from January 22, 2020 to April 29, 2020, with an overall observed case-fatality rate of 5.8%. Spatial variations were observed for both COVID-19 death outcomes and long-term ambient air pollutant levels. County-level average NO_2_ concentrations were positively associated with both COVID-19 case-fatality rate and mortality rate in single-, bi-, and tri-pollutant models (p-values<0.05). Per inter-quartile range (IQR) increase in NO_2_ (4.6 ppb), COVID-19 case-fatality rate and mortality rate were associated with an increase of 7.1% (95% CI 1.2% to 13.4%) and 11.2% (95% CI 3.4% to 19.5%), respectively. We did not observe significant associations between long-term exposures to PM_2.5_ or O_3_ and COVID-19 death outcomes (p-values>0.05), although per IQR increase in PM_2.5_ (3.4 ug/m^3^) was marginally associated with 10.8% (95% CI: −1.1% to 24.1%) increase in COVID-19 mortality rate.

**Discussions and Conclusions:** Long-term exposure to NO_2_, which largely arises from urban combustion sources such as traffic, may enhance susceptibility to severe COVID-19 outcomes, independent of longterm PM_2.5_ and O_3_ exposure. The results support targeted public health actions to protect residents from COVID-19 in heavily polluted regions with historically high NO_2_ levels. Moreover, continuation of current efforts to lower traffic emissions and ambient air pollution levels may be an important component of reducing population-level risk of COVID-19 deaths.

## Introduction

The novel human coronavirus disease 2019 (COVID-19) is an emerging infectious disease caused by severe acute respiratory syndrome coronavirus 2 (SARS-CoV-2)^1^. First identified in 2019 in Wuhan, the capital of Hubei Province, China, the COVID-19 pandemic has since rapidly spread globally. As of April 29, 2020, there have been 1,027,799 cases and 58,489 deaths confirmed in the United States^2^. Despite substantial public health efforts, the observed COVID-19 case-fatality rate (i.e. the ratio of the number of COVID-19 deaths over the number of cases) in the US is estimated to be 5.8%^3^. Although knowledge concerning the etiology of COVID-19-related disease has grown since the outbreak was first identified, there is still considerable uncertainty concerning its pathogenesis, as well as factors contributing to heterogeneity in disease severity around the globe. Environmental factors, such as urban air pollution, may play an important role in increasing susceptibility to severe outcomes of COVID-19. The impact of ambient air pollution on excess morbidity and mortality has been well-established over several decades^4-8^. In particular, major ubiquitous ambient air pollutants, including fine particulate matter (PM_2.5_), nitrogen dioxide (NO_2_), and ozone (O_3_), may have both direct and indirect systemic impact on the human body by enhancing oxidative stress and inflammation, eventually leading to respiratory, cardiovascular, and immune system dysfunction and deterioration^9-13^.

While the epidemiologic evidence is limited, previous findings on the outbreak of severe acute respiratory syndrome (SARS), the most closely related human coronavirus disease to COVID-19, revealed positive associations between air pollution and SARS case-fatality rate in the Chinese population^14^. An analysis of 213 cities in China recently demonstrated that temporal increases in COVID-19 cases was associated with short-term variations in ambient air pollution^15^. Hence, it is plausible that prolonged exposure to air pollution may have a detrimental effect on the prognosis of patients affected by COVID-19^16^. As is usual in the early literature on emerging hazards, questions remain concerning the generalizability and reproducibility of these finding, due to the lack of control for the epidemic stage-of-disease, population mobility, residual spatial correlation, and potential confounding by co-pollutants.

To address these analytical gaps and contribute towards a more complete understanding of the impact of long-term exposures to ambient air pollution on COVID-19-related health consequences, we conducted a nationwide study in the USA (3,122 counties) examining associations between multiple key ambient air pollutants, NO_2_, PM_2.5_, and O_3_, and COVID-19 case-fatality and mortality rates in both single and multi-pollutant models, with comprehensive covariate adjustment. We hypothesized that residents living in areas with higher long-term ambient air pollution levels may be more susceptible to COVID-19 severe outcomes, thus resulting in higher COVID-19 case-fatality rates and mortality rates among more heavily polluted counties.

## Methods

We obtained the number of daily county-level COVID-19 confirmed cases and deaths that occurred from January 22, 2020, the day of first confirmed case in the US, through April 29, 2020 in the US from three databases: the New York Times^17^, USAFACTS^18^, and 1Point3Acres.com^19^. In this analysis, the main COVID-19 death outcomes included two measures, the county-level COVID-19 case-fatality rate and mortality rate. The COVID-19 case-fatality rate was calculated by dividing the number of deaths over the number of people diagnosed for each US county with at least 1 or more confirmed case, which can imply the biological susceptibility towards server COVID-19 outcomes (i.e. death). The COVID-19 mortality rate was the number of COVID-19 deaths per million population, and it can reflect the severity of the COVID-19 deaths in the general population.

Three major criteria ambient air pollutants were included in the analysis, including NO_2_, a traffic-related air pollutant and a major component of urban smog, PM_2.5_, a heterogeneous mixture of fine particles in the air, and O_3_, a common secondary air pollutant^20^. We recently estimated daily ambient NO_2_, PM_2.5_, and O_3_ levels at 1 km^2^ spatial resolution across the Contiguous US using an ensemble machine learning model^21,22^. We calculated the daily average for each county based on all covered 1 km^2^ grid cells, and then further calculated the annual mean (2010-2016) for NO_2_ and PM_2.5_, and the warm-season mean (2010-2016) for O_3_, defined as May 1 to October 31. Although more recent exposure data were not available, county-specific mean concentrations of air pollutants across years are highly correlated^23^.

We compiled county-level information for several covariates that may also contribute to heterogeneity in the observed COVID-19 rates and thus may confound associations with long-term air pollution exposure. Healthcare capacity was measured by the number of intensive care unit (ICU) beds, hospital beds, and active medical doctors per 1000 people^24^. Population travel mobility index, based on anonymized location data from smartphones, was used to account for changes in travel distance in reaction to the COVID-19 pandemic^25,26^. Socioeconomic status (SES) was measured by social deprivation index^27^, a commonly used measure of area-level SES, composed of income, education, employment, housing, household characteristics, transportation, and demographics^28^. Sociodemographic covariates included population density, percentage of elderly (age ≥ 60), and percentage of male^24^. We also obtained behavioral risk factors including population mean body mass index (BMI) and smoking rate^24^, and meteorological variables^29^ including air temperature and relative humidity. Additional information about these covariates, including data sources, are given in the Technical Appendix.

## Statistical methods

We fit zero-inflated negative binomial mixed models (ZINB) to estimate the associations between longterm exposure to NO_2_, PM_2.5_, and O_3_ and COVID-19 case-fatality rates and mortality rates. The ZINB model comprises a negative binomial log-linear count model and a logit model for predicting excess zeros^30,31^. The former was used to describe the associations between air pollutants and COVID-19 case-fatality rate among counties with at least one reported COVID-19 case. The latter can account for excess zeros in counties that have not observed a COVID-19 death as of April 29, 2020. We fit single-pollutant, bi-pollutant, and tri-pollutant models, in order to estimate the effects of each pollutant without and with control for co-pollutants. All analyses were conducted at the county level. For the negative binomial count component, results are presented as percent change in case-fatality rate or mortality rate per interquartile range (IQR) increase in each air pollutant concentration. IQRs were calculated based on mean air pollutant levels across all 3,122 counties. Similar results are presented as odds ratios for the excess zero component. We included a random intercept for each state because observations within the same state tended to be correlated, potentially due to similar COVID-19 responses, quarantine and testing policies, healthcare capacity, sociodemographic, and meteorological conditions.

As different testing practices may bias outcome ascertainment, we adjusted for state-level COVID-19 test positive rate (i.e. a high positive rate might imply that the confirmed case numbers were limited by the ability of testing, thus upward-biasing the case-fatality). To model how different counties may be at different time points of the epidemic curve (i.e., phase-of-epidemic), we adjusted for days both since the first case and since the 100^th^ case within a county through April 29. In addition, we adjusted for potential confounders including county-level healthcare capacity, population mobility, sociodemographic, SES, behavior risk factors, and meteorological factors, as described above.

To control for potential residual spatial trends and confounding, we included spatial smoothers within the model using natural cubic splines with 5 degrees freedom for both county centroid latitude and longitude. To examine the presence of spatial autocorrelation in the residuals, we calculated Moran’s I of the standardized residuals of tri-pollutant main models among counties within each state. Statistical tests were 2-sided and statistical significance and confidence intervals were calculated with an alpha of 0.05. All statistical analyses were conducted used R version 3.4.

## Sensitivity analyses

We conducted a series of sensitivity analyses to test the robustness of our results to outliers, confounding adjustment, and epidemic timing (Supplementary Appendix Figures S1 and S2). Given that New York City has far higher COVID-19 cases and deaths than any other region, we excluded all five counties within New York City in one sensitivity analysis. In another, we restricted the study to the most recent 4 weeks (April 1 to April 29), when the case count and death count may be more reliable and accurate compared to earlier periods. We also conducted sensitivity analysis by using air pollution data averaged between 2000 to 2016. To assess the importance of individual confounder covariates, we fit models by omitting a different set of covariates for each model iteration and compared effect estimates.

## Results

A total of 3,122 US counties were considered in the current analysis, with confirmed cases reported in 2,810 (90.0%) and deaths in 1,443 (46.2%). By April 29, 2020, 1,027,799 COVID-19 cases and 58,489 deaths were reported nationwide (Table 1). Among the counties with at least one reported COVID-19 case, the average county-level case-fatality rate was 3.7 ± 7.0% (mean ± standard deviation), and the average mortality rate was 69.4 ± 183.1 per 1 million people. Spatial variations were observed on COVID-19 case-fatality and mortality rates, where Michigan had the highest average county-level case-fatality rate of 8.2% and New York had the largest mortality rate of 1,117.2 deaths per 1 million people. The lowest case-fatality rate and mortality rate were observed in Wyoming (0.7%) and North Dakota (11.8 deaths per million people), respectively (Figure 1).

**Table 1.**
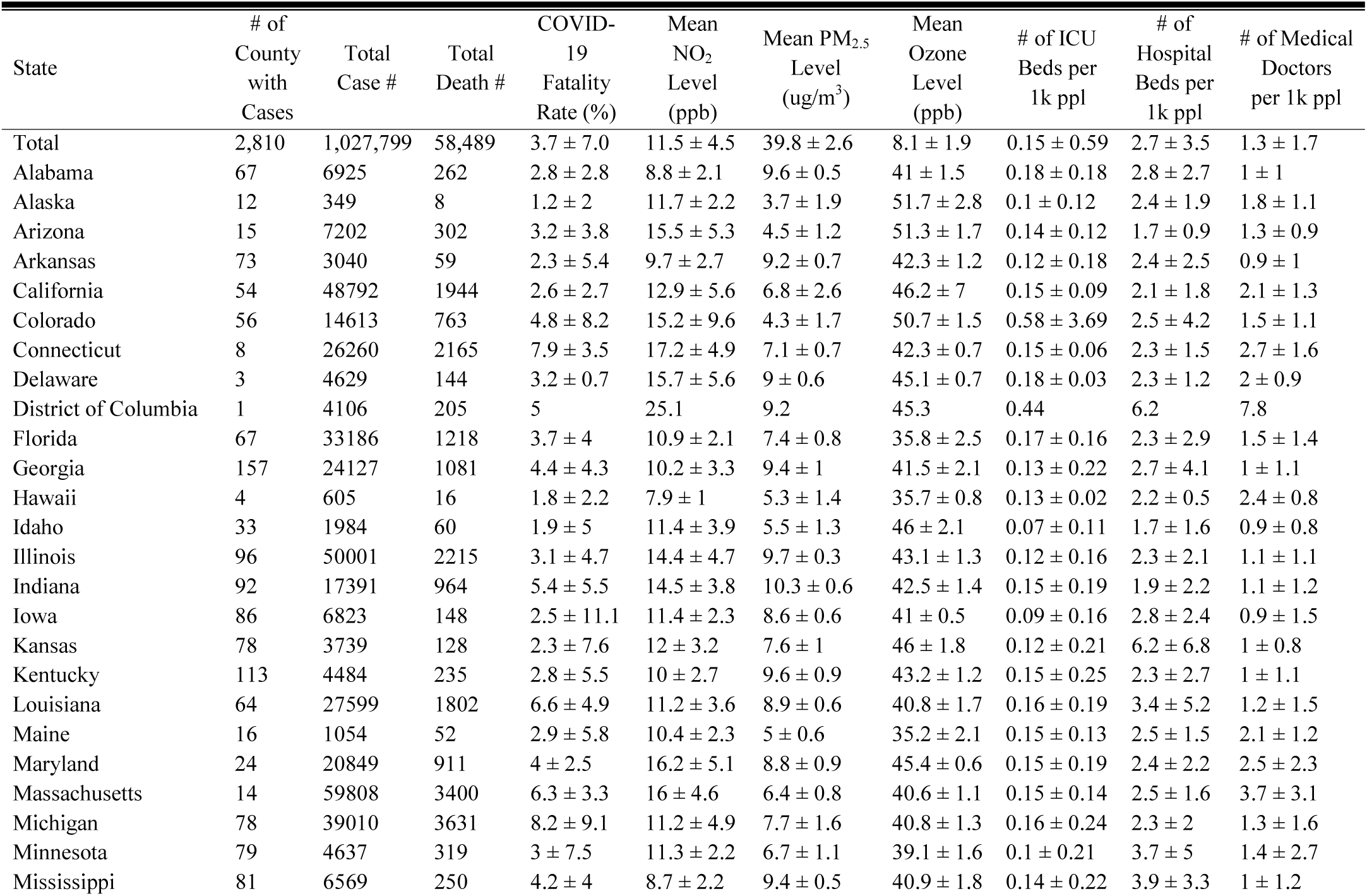
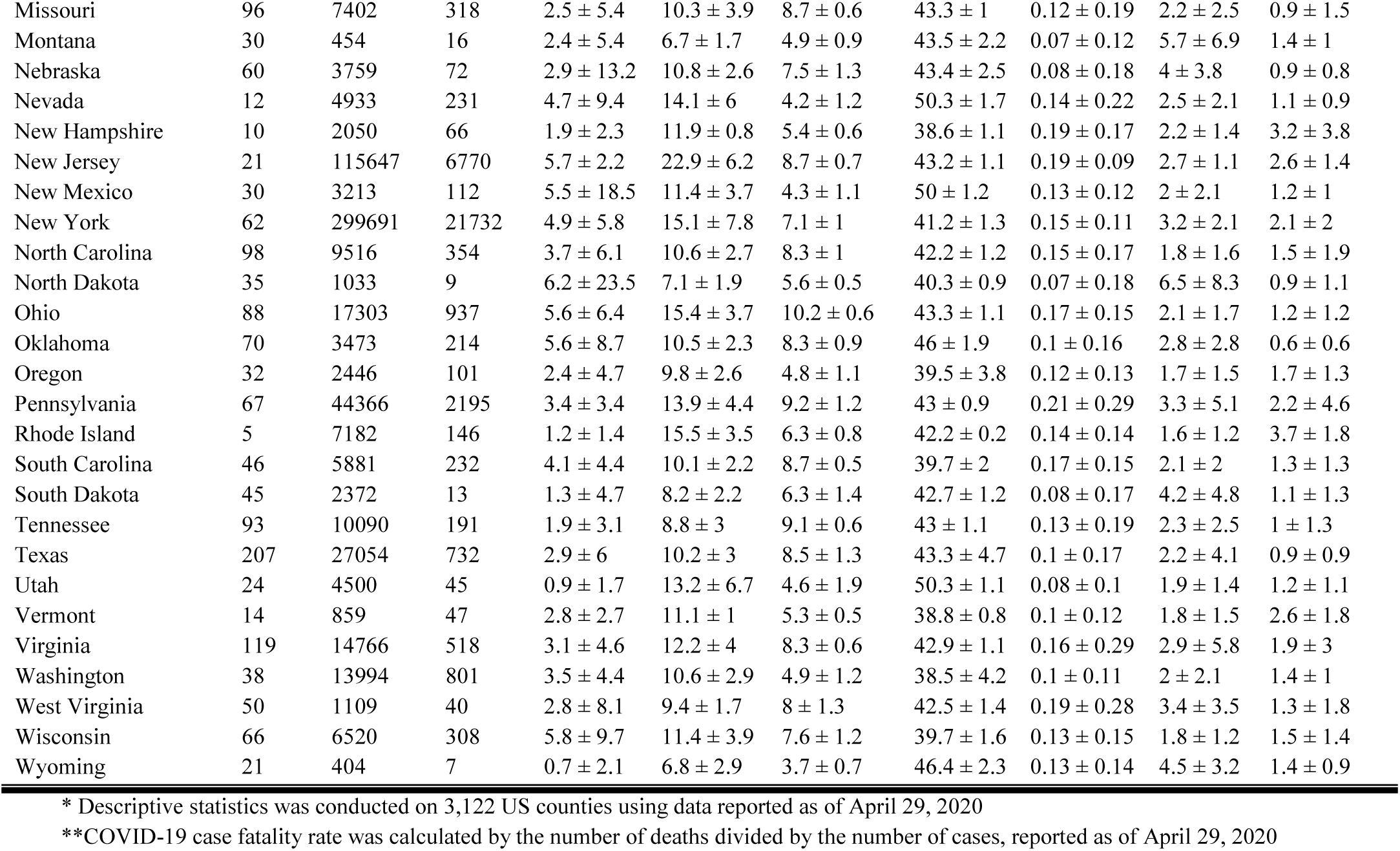
Descriptive statistics* was conducted on 3,122 US counties using data reported as of April 29, 2020.

**Figure 1.**
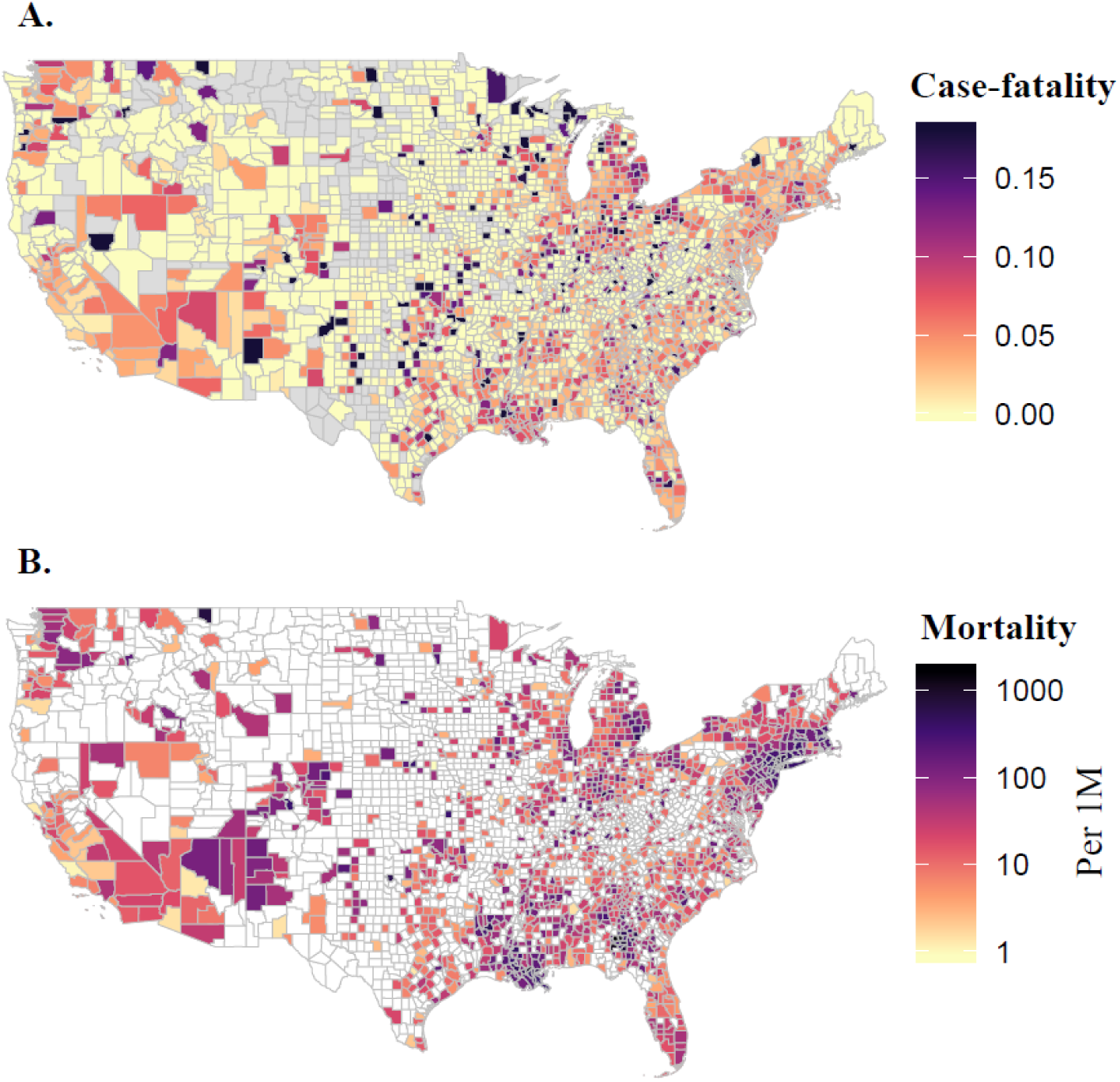
County-level COVID-19 Case-fatality Rate (A) and Mortality Rate per 1 Million people (B) as of April 29, 2020.

Long-term (2010-2016) average concentrations across the contiguous US ranged from 5.8 to 19.3 parts per billion (ppb; 5^th^ and 95^th^ percentiles, respectively) NO_2_, 3.8 to 10.4 (μg/m^3^ PM_2.5_, and 37.2 to 49.7 ppb for warm-season average ozone concentrations, respectively (Figure 2). The highest NO_2_ levels were in New York, New Jersey, and Colorado, and the lowest in Montana, Wyoming, and South Dakota. California and Pennsylvania had the highest PM_2.5_ concentrations, and the highest O_3_ levels were in Colorado, Utah, and California.

**Figure 2.**
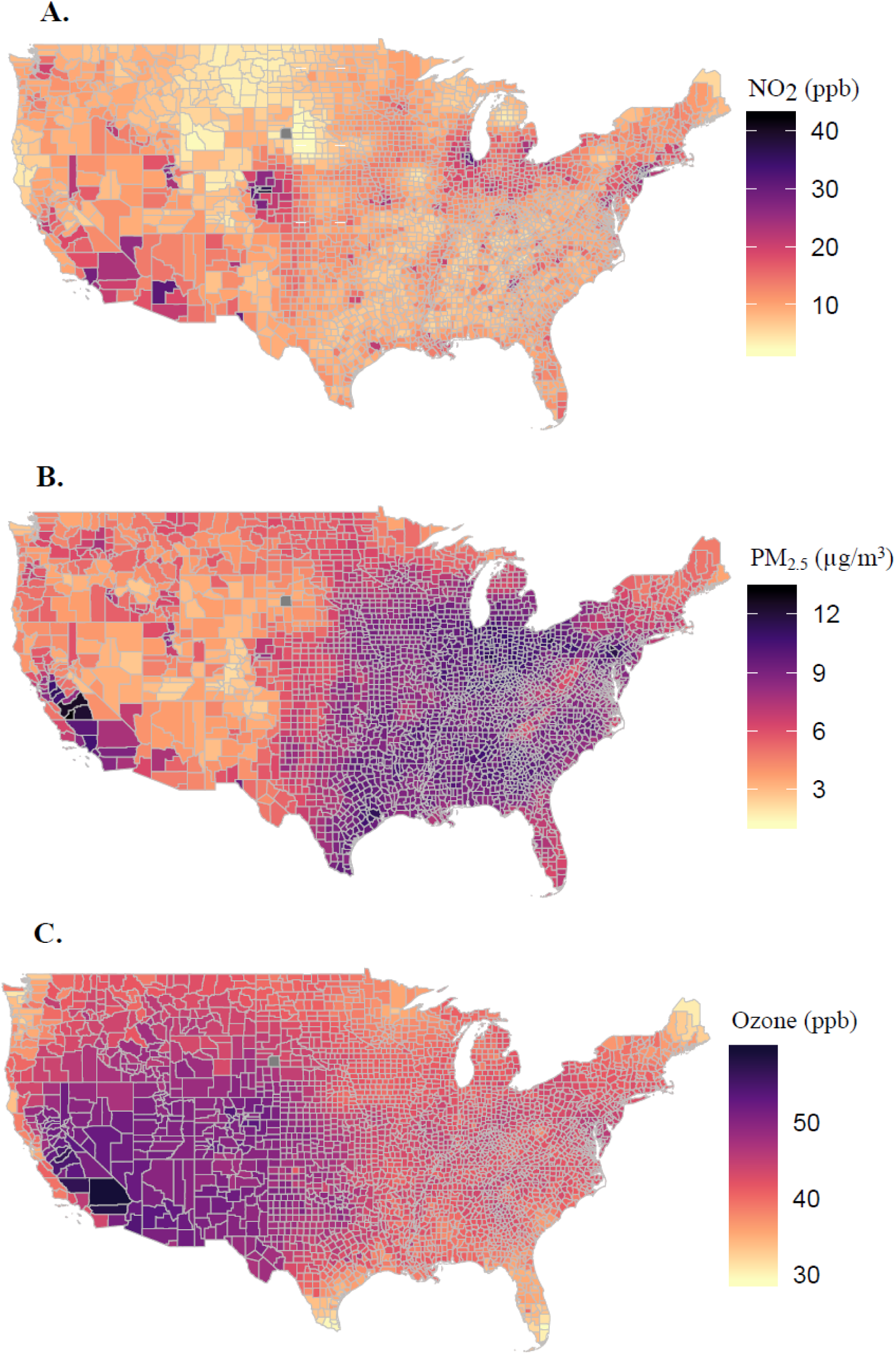
County-level Annual Average Concentrations of Nitrogen Dioxide (NO_2_, A), Fine Particulate Matter (PM_2.5_, B), and Ozone (C) for the period 2010-2016.

We observed significant positive associations between NO_2_ levels and both county-level COVID-19 case-fatality rate and mortality rate (p=0.02 and p<0.001, respectively, Table S1 and Figure 3), when controlling for covariates. In single pollutant models, COVID-19 case-fatality and mortality rates were associated with increases of 7.1% (95% CI: 1.2% to 13.4%) and 11.2% (95% CI: 3.4% to 19.5%), respectively, per IQR (~4.6 ppb) increases in NO_2_. These results imply that one IQR reduction in long-term exposure to NO_2_ level would have avoided 4,181 deaths (95% CI: 718 to 7,845) among those tested positive for the virus and 19.9 deaths (95% CI: 6.1 to 34.8) per million people in the general population, as of April 29, 2020.

**Figure 3.**
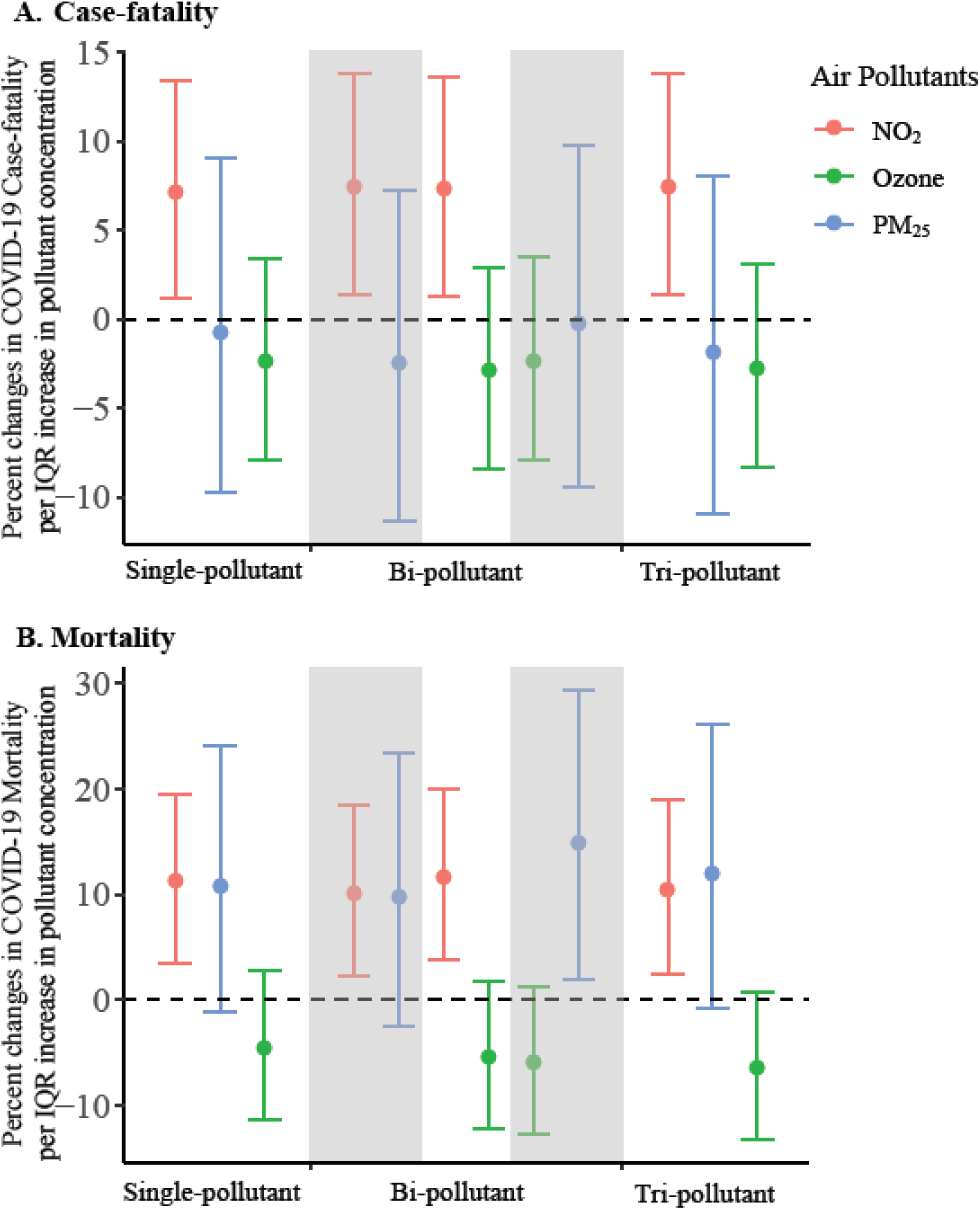
Percent Change in County-level COVID-19 Case-fatality Rate (A) and Mortality Rate (B) Per Inter Quartile Range (IQR) increase in Long-Term Air Pollutant Concentrations. Effect estimates and 95% confidence interval were calculated using county-level concentrations of nitrogen dioxide (NO_2_, red), ozone (green), and fine particulate matter (PM_2.5_, blue) averaged between 2010-2016, controlling for covariates including county-level number of case per 1000 people, social deprivation index, population density, percent of residents over 60 years old, percent of male, body mass index, smoking rate, number of regular hospital beds per 1000 people, number of intensive units beds per 1000 people, number of medical doctors per 1000 people, average mobility index assessed in March and April 2020, average temperature and humidity between January 22 to April 29, 2020, state-level COVID-19 test positive rate as of April 29, 2020, and spatial smoother with a degree freedom of 5 for both latitude and longitude. IQRs of NO_2_, PM_2.5_, and O_3_ averaged between 2010-2016 were 4.6 parts per billion (ppb), 3.4 ug/m^3^, and 2.8 ppb, respectively.

Results from the zero-component model agrees with the count model where higher NO_2_ concentrations were associated with a decrease in the probability of observing zero COVID-19 death (Appendix Table S1). The strength and magnitude of the associations between NO_2_ and both COVID-19 case-fatality rate and mortality rate persisted across single, bi-, and tri-pollutant models (Figure 3).

In contrast, PM_2.5_ was not associated with COVID-19 case-fatality rate (p=0.87) but was marginally associated with higher COVID-19 mortality rate (p=0.08), where one IQR (3.4 ug/m^3^) increase in PM_2.5_ was associated with 10.8% (95% CI: −1.1% to 24.1%) increase in COVID-19 mortality rate. Null associations were found between long-term exposure to O_3_ and both COVID-19 case-fatality and mortality rates (p=0.42 and p=0.22, respectively). The Moran’s I and p-values (Appendix Table S2) from these models suggested that most spatial correlation in the data has been accounted for.

Results remained robust and consistent across 30 sets of sensitivity analyses (Appendix Figures S1 and S2). When we restricted the analyses to data reported between April 1 to April 29, when COVID-19 tests were more readily available, significant associations remained between NO_2_ and COVID-19 case-fatality and mortality rates, and no associations were found with PM_2.5_ or O_3_. We also observed similar trends pointing to associations with NO_2_ when excluding the New York City. In addition, even though 679 counties (22%) had missing behavioral risk data, the analysis omitting behavioral risk factors yielded similar results.

## Discussion

In this nationwide study, we used county-level information on long-term air pollution and corresponding health, behavioral, and demographic data to examine associations between long-term exposures to key ambient air pollutants and COVID-19 death outcomes in both single and multi-pollutant models. We observed significant positive associations between NO_2_ levels and both county-level COVID-19 case-fatality rate and mortality rate, while null associations were found for long-term PM_2.5_ and O_3_ exposures. These results provide additional initial support for the interpretation that long-term exposure to NO_2_, a component of urban air pollution related to traffic, may enhance susceptibility to severe COVID-19 outcomes. These findings may help identify susceptible and high-risk populations, especially those living in areas with historically high NO_2_ pollution, including the metropolitan areas in New York, New Jersey, Colorado, and Michigan. Given the rapid escalation of COVID-19 spread and associated mortality in the US, swift and coordinated public health actions, including strengthened enforcement on social distancing and expanding healthcare capacity, are needed to protect these and the other vulnerable populations. Although average NO_2_ concentrations have decreased gradually over the past decades, it is critical to continue enforcing air pollution regulations to protect public health, given that health effects occur even at very low concentrations^8^.

Among the sparse studies on the link between air pollution and COVID-19, our findings are consistent with a recent European study that reported 78% of the COVID-19 deaths across 66 administrative regions in Italy, Spain, France and Germany, occurred in the five most polluted regions with the highest NO_2_ levels^32^. Another recent paper reported correlations between high levels of air pollution and high death rates seen in northern Italy^33^. However, major questions remain concerning the robustness and generalizability of these early findings, due to the lack of control for population mobility, multipollutant exposures, and most importantly, potential residual spatial autocorrelation.

With the current analysis, we hopefully contribute to the questions involving the potential link between urban air pollution and COVID-19 risk in addressing these issues. We examined two major COVID-19 death outcomes, the county-level case-fatality rate and the mortality rate. The case-fatality rate can imply on the biological susceptibility towards server COVID-19 outcomes (i.e. death), while the mortality rate can offer information of the severity of the COVID-19 deaths in the general population. Looking at both case-fatality and mortality will contribute to a more comprehensive understanding on the impact of air pollution exposures on COVID-19 death outcomes. Our study included an assessment of three major air pollutants using high spatial resolution maps, the use of recent county-level data, consideration for both single and multi-pollutant models, and control of county-level mobility. Given that the stage of the COVID-19 epidemic might depend on the size and urbanicity of the county, we included the time of the first and 100^th^ case for each county in the models as covariates to minimize the possibility that the observed associations are confounded by epidemic timing due to unmeasured location and population-level characteristics. Due to the cross-sectional design, we controlled for potential spatial trends by including flexible spatial trends in the main analysis, and evaluated residual autocorrelation using Moran’s I statistic. Our analyses indicated that the presence of spatial confounding was substantial, necessitating the use of spatial smoothing. We observed statistically significant PM_2.5_ and O_3_ associations with COVID-19 mortality in models without controlling for spatial smoothers (Appendix Figures S1 and S2). Finally, we conducted a total of 30 sets of sensitivity analyses and observed robust and consistent results.

Although the various social distancing measures around the US have reduced vehicle traffic and urban air pollution, it is plausible that long-term exposure to urban air pollutants like NO_2_ may have direct and indirect effects within the human body, making people more biologically susceptible to severe COVID-19 outcomes. NO_2_ can be emitted directly from combustion sources or produced from the titration of NO with O_3_. NO_2_ and nitric oxide (NO) have relatively short atmospheric lifetime, thus having larger spatial heterogeneity compared to more regionally distributed pollutants such as PM_2.5_ and O_3_. As a result, the spatial distribution of NO_2_ represents the intensity of anthropogenic activity, especially emissions from traffic and power plants. As a reactive free radical, NO_2_ plays a key role in photochemical reactions that produce other secondary pollutants, including ozone and secondary particulate matter. In our analysis of three major air pollutants, however, NO_2_ showed strong and independent effects with COVID-19 case-fatality rate and mortality, meaning that the effects of NO_2_ may not be mediated by PM_2.5_ and O_3_. Even so, we cannot rule out the possibility that NO_2_ is serving as a proxy for other traffic-related air pollutants, such as soot, trace metals, or ultrafine particles. Long-term exposures to NO_2_ have been associated with acute and chronic respiratory diseases, including increased bronchial hyperresponsiveness, decreased lung function, and increased risk of respiratory infection and mortality^34-36^. In addition, as a highly reactive exogeneous oxidant, NO_2_ can induce inflammation and enhance oxidative stress, generating reactive oxygen and nitrogen species, which may eventually deteriorate the cardiovascular and immune systems^37,38^. The impact of long-term exposure to PM_2.5_ on excess morbidity and mortality has also been well-established^4-5,8^. An early unpublished report that explored the impacts of air pollution on mortality found that 1 μg/m^3^ PM_2.5_ was associated with 8% increase in COVID-19 mortality rates in the USA^39^. The study was conducted in a single pollutant model and did not investigate on COVID-19 case-fatality rates. In contrast, we found only marginally significant associations between COVID-19 mortality rates and PM_2.5_. Specifically, the magnitude and strength of this association observed in the current analysis were weaker, mainly due to our control of the spatial trends and residual autocorrelation, which may have confounded the previous study findings^34^. In addition, PM_2.5_ was not associated with COVID-19 case-fatality rate across all single and multipollutant models, indicating that it may have less impact on biological susceptibility to severe COVID-19 outcomes compared to NO_2_.

We acknowledge that our study is limited in several key areas. First, the cross-sectional study design reduced our ability to exploit temporal variation and trends in COVID-19 deaths, an important determinant in establishing causal inference. Towards this end, time-series analyses of air pollution and COVID-19 case-fatality rates and corresponding mortality rates will be important; these data will only be available in the future. Second, actual death counts are likely underestimated, particularly during the early stages of the outbreak, with highly dynamic reported fatality rates, increasing from 1.8% to 5.8% in the past two months. However, results using data from only the most recent four weeks were largely unchanged, suggesting that differential errors in reporting or testing for COVID-19 may not have exerted much influence on these findings. Third, although we controlled for many potential confounders such as population density, we cannot rule out the possibility that NO_2_ might be a proxy of urbanicity. The exclusion of climate meteorological variables and SES – two factors that have received substantial attention regarding the outbreak – did not alter the main results. In addition, testing data only came from state or federal agencies, while tests from private labs were not available. Due to the lack of county-level data, we could not account for the percentage of hospitalized cases or ICU use among cases or deaths, the number of available ventilators, and the underlying health conditions of cases likely to increase death risk (e.g., chronic obstructive pulmonary disease). Also, as a classic traffic related air pollutant, NO_2_ can exhibit spatial variation within a county, which may not be captured in our analysis. Identification of NO_2_ pollution hotspots within a county may be warranted.

## Conclusions

We found statistically significant, positive associations between long-term exposure to NO_2_ and COVID-19 case-fatality rate and mortality rate, independent of PM_2.5_ and O_3_. Prolonged exposure to this urban traffic-related air pollutant may be an important risk factor of severe COVID-19 outcomes. The results support targeted public health actions to protect residents from COVID-19 in heavily polluted regions with historically high NO_2_ levels. Moreover, continuation of current efforts to lower traffic emissions and ambient air pollution levels may be an important component of reducing population-level risk of COVID-19 deaths.

## Data Availability

Data will be made available when requested

## Author Contribution

D.L., L.S., and H.C. designed research and directed its implementation; D.L., L.S., J.Z., P.L., J.S., and S.G. prepared datasets; D.L., H.C., and L.S. analyzed data; D.L., L.S., and J.Z. wrote the paper and made the tables; L.S. made the figures; and all authors contributed to the revision of the manuscript.

## Potential Conflicts of Interest

The authors have no conflicts of interest relevant to this article to disclose

## Acknowledgement

This project was supported by Emory HERCULES exposome center through the National Institute of Environmental Health Sciences (grant number P30ES019776). S.G. acknowledges the funding support provided by the National Science Foundation (Award No. BCS-2027375). Any opinions, findings, and conclusions or recommendations expressed in this material are those of the author(s) and do not necessarily reflect the views of the National Institutes of Health and the National Science Foundation. The authors would like to thank Qian Di, Weeberb J. Requia, and Yagung Wei for contribution to the generation of air pollution data.

## Data Sharing Statement

Data will be made available when requested.

## Supplementary Appendix for

### Supplementary Tables and Figures

**Table S1.** Model Effect Estimates on Zero-inflated Negative Binomial Mixed Models to Examine the Associations between Long-term Exposure to Air Pollution and COVID-19 case-fatality rate or mortality.

| Pollutant | COVID-19 Case-Fatality |  |  | COVID-19 Mortality |  |  |
| --- | --- | --- | --- | --- | --- | --- |
|  | Main Effect Estimate* | 95% Confidence Interval | p-values | Main Effect Estimate | 95% Confidence Interval | p-values |
| **Single Pollutant Model—Non-zero Components** |  |  |  |  |  |  |
| NO<sub>2</sub> | 1.015 | 1.003 to 1.028 | 0.02 | 1.023 | 1.007 to 1.039 | <0.001 |
| PM<sub>2.5</sub> | 0.996 | 0.953 to 1.041 | 0.87 | 1.049 | 0.995 to 1.107 | 0.08 |
| O<sub>3</sub> | 0.993 | 0.975 to 1.010 | 0.42 | 0.986 | 0.964 to 1.008 | 0.22 |
| **3- Pollutant Model—Non-zero Components** |  |  |  |  |  |  |
| NO<sub>2</sub> | 1.016 | 1.003 to 1.029 | 0.02 | 1.022 | 1.005 to 1.038 | <0.001 |
| PM<sub>2.5</sub> | 0.991 | 0.947 to 1.037 | 0.70 | 1.054 | 0.996 to 1.115 | 0.07 |
| O<sub>3</sub> | 0.992 | 0.974 to 1.010 | 0.36 | 0.979 | 0.957 to 1.002 | 0.08 |
| **Single Pollutant Model—Zero Components** |  |  |  |  |  |  |
| NO<sub>2</sub> | 0.963 | 0.938 to 0.988 | <0.001 | 0.943 | 0.917 to 0.969 | <0.001 |
| PM<sub>2.5</sub> | 0.843 | 0.779 to 0.912 | <0.001 | 0.689 | 0.630 to 0.754 | <0.001 |
| O<sub>3</sub> | 0.860 | 0.828 to 0.892 | <0.001 | 0.792 | 0.760 to 0.825 | <0.001 |
| **3-Pollutant Model—Zero Components** |  |  |  |  |  |  |
| NO<sub>2</sub> | 0.969 | 0.945 to 0.994 | 0.02 | 0.949 | 0.922 to 0.978 | <0.001 |
| PM<sub>2.5</sub> | 0.902 | 0.831 to 0.980 | 0.01 | 0.772 | 0.703 to 0.848 | <0.001 |
| O<sub>3</sub> | 0.869 | 0.838 to 0.902 | <0.001 | 0.804 | 0.772 to 0.838 | <0.001 |

\*Effect estimate based on per unit increase in air pollutants

**Table S2.** Moran’s I test for spatial autocorrelation in residuals from tri-pollutant models for COVID-19 Case-fatality Rate and Mortality Rate for each US state.

|  | **Case-fatality Rate** |  | **Mortality Rate** |  |
| --- | --- | --- | --- | --- |
| **State** | **Moran's I** | ***p*-value** | **Moran's I** | ***p*-value** |
| **Alabama** | 0.023005 | 0.666791 | -0.016454 | 0.997143 |
| **Arizona** | -0.069966 | 0.971086 | 0.159086 | 0.183565 |
| **Arkansas** | -0.045066 | 0.776283 | 0.045421 | 0.548356 |
| **California** | 0.024098 | 0.498309 | 0.058227 | 0.219995 |
| **Colorado** | -0.019308 | 0.900768 | -0.069735 | 0.60085 |
| **Connecticut** | -0.249311 | 0.733068 | -0.213497 | 0.835009 |
| **Delaware** | -0.338612 | 1 | NA | NA |
| **District of Columbia** | NA | NA | NA | NA |
| **Florida** | -0.033006 | 0.771992 | -0.046262 | 0.617719 |
| **Georgia** | 0.056318 | 0.172308 | 0.051514 | 0.200098 |
| **Idaho** | -0.053368 | 0.95988 | -0.003237 | 0.717008 |
| **Illinois** | 0.056139 | 0.412049 | 0.096366 | 0.177248 |
| **Indiana** | -0.116814 | 0.169918 | 0.172651 | 0.02579 |
| **Iowa** | -0.054103 | 0.568083 | -0.027161 | 0.8795 |
| **Kansas** | 0.07953 | 0.061874 | -0.03842 | 0.901546 |
| **Kentucky** | -0.047995 | 0.623543 | 0.062124 | 0.36778 |
| **Louisiana** | -0.029227 | 0.902967 | 0.035875 | 0.539011 |
| **Maine** | -0.135679 | 0.44648 | -0.141616 | 0.384822 |
| **Maryland** | 0.028251 | 0.611963 | 0.236883 | 0.041622 |
| **Massachusetts** | -0.041547 | 0.826158 | -0.096154 | 0.947948 |
| **Michigan** | -0.03011 | 0.730351 | 0.038998 | 0.252498 |
| **Minnesota** | 0.019843 | 0.417719 | -0.01777 | 0.986115 |
| **Mississippi** | 0.096789 | 0.043456 | 0.044718 | 0.271883 |
| **Missouri** | 0.030381 | 0.500362 | 0.073753 | 0.192159 |
| **Montana** | 0.052994 | 0.060993 | 0.076018 | 0.031501 |
| **Nebraska** | -0.040916 | 0.86608 | -0.059581 | 0.722833 |
| **Nevada** | 0.337241 | 0.038866 | 0.38327 | 0.035115 |
| **New Hampshire** | 0.131492 | 0.042746 | 0.103783 | 0.081307 |
| **New Jersey** | 0.175217 | 0.087009 | 0.126441 | 0.178484 |
| **New Mexico** | -0.045416 | 0.979142 | 0.153729 | 0.258981 |
| **New York** | 0.013377 | 0.579773 | -0.033898 | 0.807794 |
| **North Carolina** | -0.050907 | 0.34774 | -0.032161 | 0.647075 |
| **North Dakota** | 0.109892 | 0.49598 | 0.354751 | 0.100111 |
| **Ohio** | -0.041737 | 0.707563 | 0.107715 | 0.139459 |
| **Oklahoma** | 0.06193 | 0.010202 | 0.102011 | 0.000142 |
| **Oregon** | 0.007053 | 0.350889 | 0.019009 | 0.24168 |
| **Pennsylvania** | 0.108983 | 0.133666 | 0.348912 | 0.000011 |
| **Rhode Island** | -0.256215 | 0.973717 | -0.225587 | 0.897816 |
| **South Carolina** | 0.062342 | 0.49757 | 0.102716 | 0.23941 |
| **South Dakota** | -0.034629 | 0.740424 | -0.059895 | 0.913713 |
| **Tennessee** | -0.007701 | 0.958532 | 0.110452 | 0.166716 |
| **Texas** | 0.00567 | 0.215483 | -0.00599 | 0.965107 |
| **Utah** | 0.445684 | 0.000382 | 0.152707 | 0.073403 |
| **Vermont** | 0.019688 | 0.629256 | 0.269936 | 0.117271 |
| **Virginia** | 0.410102 | 0.000001 | 0.298699 | 0.000003 |
| **Washington** | 0.005076 | 0.781592 | 0.05457 | 0.507951 |
| **West Virginia** | 0.112795 | 0.000103 | 0.187581 | 0.06824 |
| **Wisconsin** | -0.039313 | 0.67382 | 0.095091 | 0.018566 |
| **Wyoming** | -0.215725 | 0.005326 | -0.148779 | 0.047632 |

**Figure S1.**
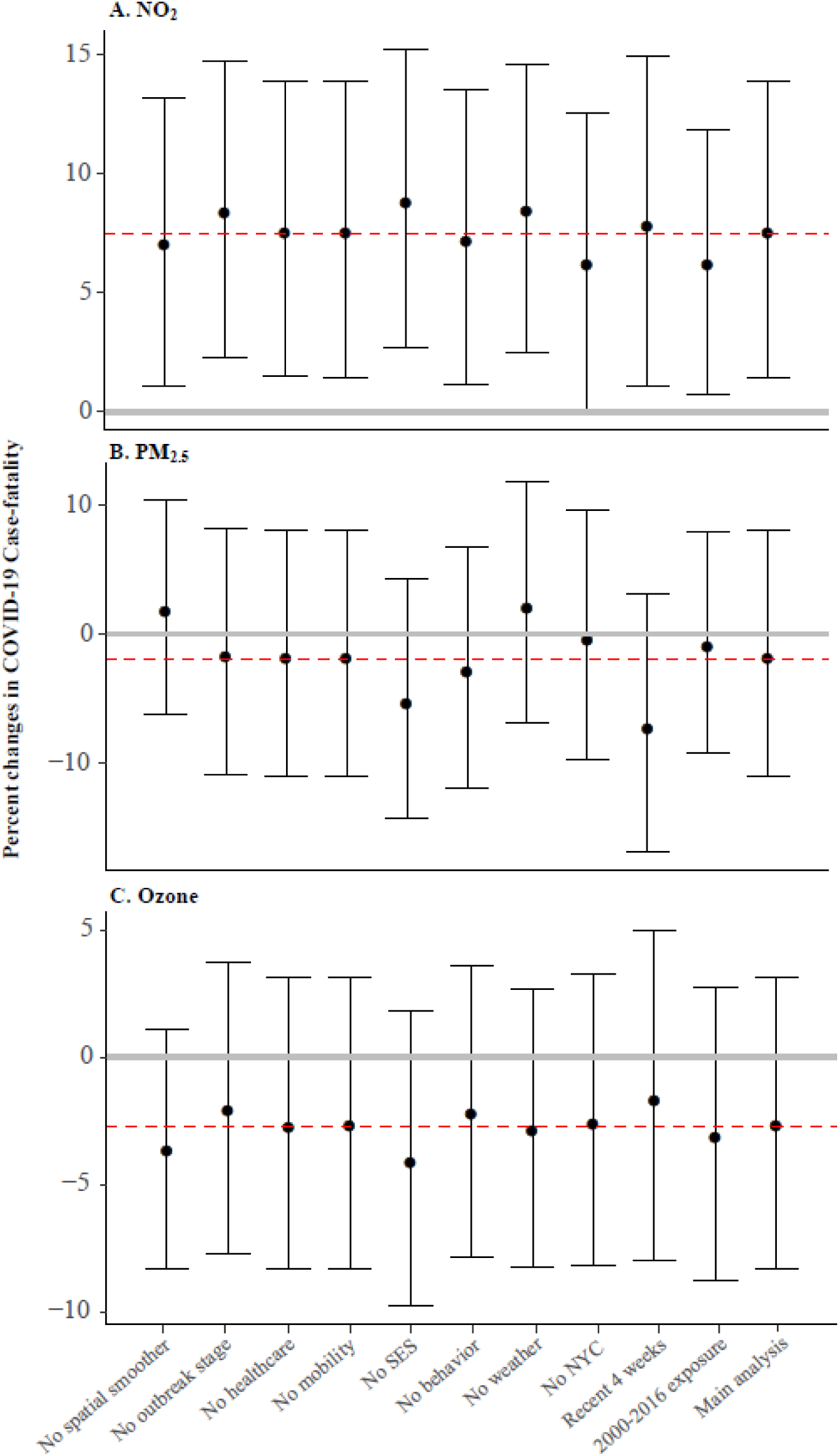
Percent Change in COVID-19 Case-fatality Rate Per Inter Quartile Range (IQR) increase in (A) NO_2_, (B) PM_2.5_, and (C) Ozone Concentrations in the Sensitivity Analysis. The red line represents the estimated effects in the main analysis. All results were derived from the tri-pollutant models.

**Figure S2.**
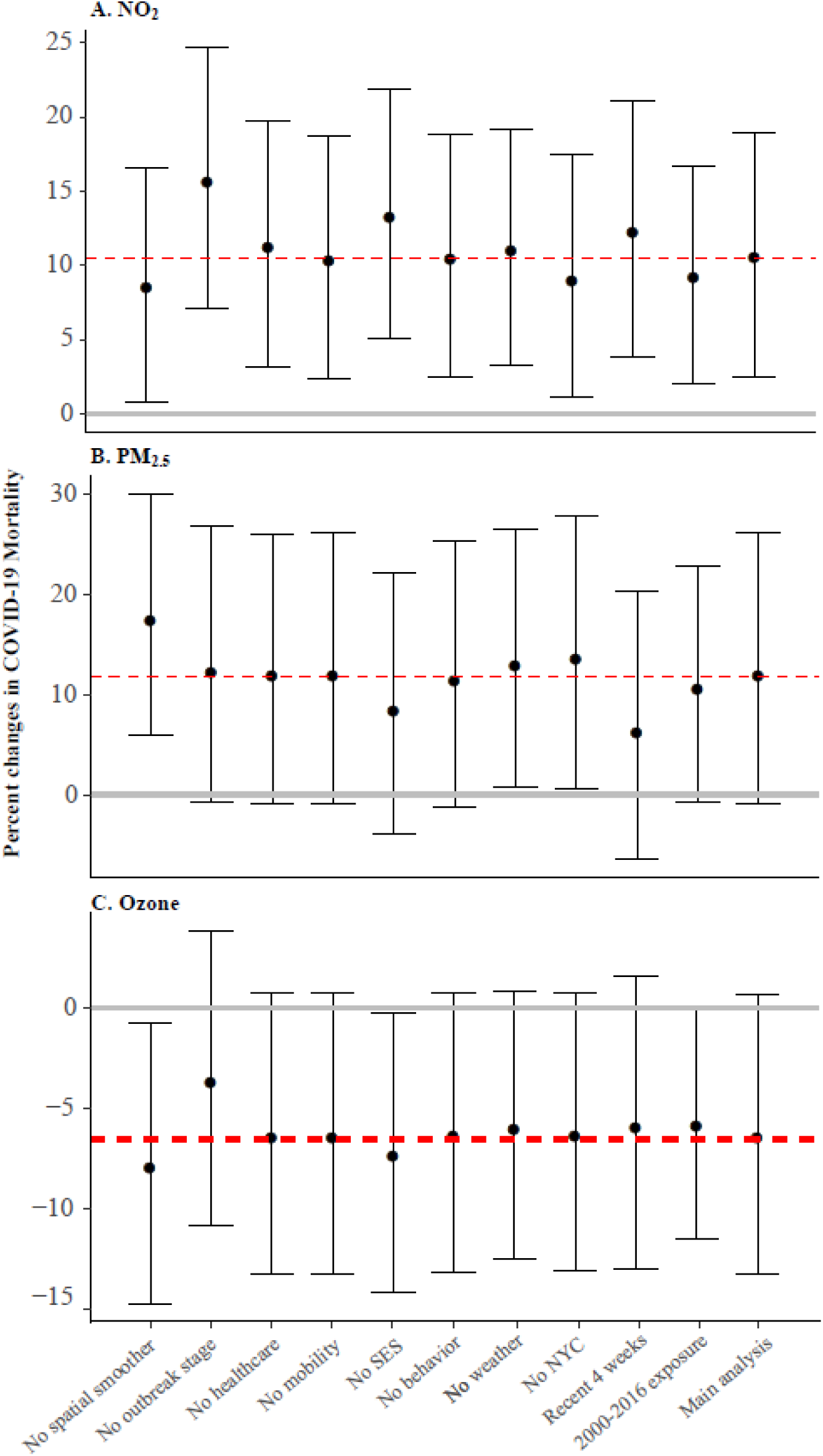
Percent Change in COVID-19 Mortality Rate Per Inter Quartile Range (IQR) Increase in (A) NO_2_, (B) PM_2.5_, and (C) Ozone Concentrations in the Sensitivity Analysis. The red line represents the estimated effects in the main analysis. All results were derived from the tri-pollutant models.

## Appendix-Technical Appendix

### COVID-19 case-fatality rate

We obtained the number of daily county-level COVID-19 confirmed cases and deaths that occurred from January 22, 2020, the day of first confirmed case in the US, through April 29, 2020 in the US from three databases: the New York Times, the USAFACTS, and 1Point3Acres.com. Each of these databases provide real-time data by retrieving information on official reports from state and local health agencies. After data acquisition from these sources, we compared the number of confirmed COVID-19 cases and deaths in each US county (identified by the Federal Information Processing Standards, FIPS code) across all databases for accuracy and consistency. In case of discrepancy, county-level case and death number were corrected by manually checking the data reported from the corresponding state and local health department websites. We calculated county-level COVID-19 case-fatality rate by dividing the number of deaths over the number of people diagnosed with COVID-19 for each US county with at least 1 or more confirmed case, as reported by April 29, 2020. Of all the data reported as of April 29, 2020, confirmed cases and deaths with unassigned counties were excluded in the analysis.

### Air pollution

Three major criteria ambient air pollutants were included in the analysis, including NO_2_, a traffic-related air pollutant and a major component of urban smog, PM_2.5_, and O_3_. We recently estimated daily ambient PM_2.5_, NO_2_, and O_3_ levels at 1 km^2^ spatial resolution across the contiguous US an ensemble machine learning model with ground measurements, satellite-data products, chemical transport model output, meteorological and land-use information as predictors^22,23^. We calculated the daily average for each county based on all covered 1 km^2^ grid cells, and then further calculated the annual mean (2010-2016) for PM_2.5_ and NO_2_ and the warm-season mean (2010-2016) for O_3_, defined as May 1 to October 31, as surrogates for long-term PM_2.5_, NO_2_, and O_3_ exposures, respectively. More recent exposure data were not available at the time of this analysis. However, county-specific mean values of an air pollutant among different years are highly correlated.

### Covariates

We compiled county-level information for several covariates that could also explain heterogeneity in the observed COVID-19 rates and may confound associations with long-term air pollution exposure. Healthcare capacity was measured by the number of intensive care unit (ICU) beds, hospital bed, and active medical doctors per 1000 people. Number of ICU beds were based on Kaiser Health News analysis of 2018 and 2019 hospital cost reports filed to the Centers for Medicare & Medicaid Services. Numbers of active medical doctors and hospital bed of 2017 were obtained from the Area Health Resources Files. Based on the number of COVID-19 tests performed in each state, we calculated a positive rate (i.e., the percentage of specimens tested that are positive for COVID-19). Travel mobility index, based on anonymized location data from smartphones, was used to account for changes in travel distance in reaction to the COVID-19 pandemic. Socioeconomic status (SES) was measured by social deprivation index, a composite measure of area-level deprivation that takes into account income, education, employment, housing, household characteristics, transportation, and demographics. Sociodemographic covariates included population density, percentage of elderly (age ≥ 60), and percentage of male. We also obtained behavioral risk factors including population mean BMI and smoking rate, and meteorological variables including air temperature and relative humidity (converted from specific humidity). All covariates were linked to the COVID-19 data using FIPS code and additional details on data source and process are given in the Supplementary Appendix.

### Statistical methods

We fit zero-inflated negative binomial mixed models (ZINB) to examine the associations between long-term exposure to PM_2.5_, NO_2_, and O_3_ and COVID-19 case-fatality rate or mortality. The ZINB model comprises a negative binomial log-linear count model and a logit model for predicting excess zeros^30,31^. The former was used to describe the associations between air pollutants and COVID-19 case-fatality rate among counties with at least one reported COVID-19 case. The latter can account for excess zeros in counties that have not observed a COVID-19 death as of April 29, 2020. We fit single-pollutant, bi-pollutant, and tri-pollutant models, with all analyses conducted at the county level. For the negative binomial count component, results are presented as percent change in case-fatality rate or mortality rate per interquartile range (IQR) increase in each air pollutant concentration. IQR was calculated on national levels. Similar results are presented as odds ratio for the excess zero component. We included a random intercept for each state because observations within the same state tend to be correlated due to similar COVID-19 responses, quarantine and testing policies, healthcare capacity, sociodemographic, and meteorological conditions.

As different testing practices may bias outcome ascertainment, we adjusted for state-level COVID-19 test positive rate (i.e. high positive rate might imply that the confirmed case numbers were limited by the ability of testing, and the case-fatality can be biased high). To model how different counties may be at different time points of the epidemic curve (i.e., phase-of-epidemic), we adjusted for days both since the first case and since the 100^th^ case (i.e., case counts reaching 100) within a county through April 29 as a measure of epidemic timing. To account for how people may have reacted to the social distancing guidelines imposed during the COVID-19 outbreak, we adjusted for county-level travel mobility index. In addition, we considered potential confounding by county-level healthcare capacity, sociodemographic, SES, behavior risk factors, and meteorological factors. Because county-specific population densities span 5 orders of magnitude, we adjusted for density using a logarithmic transformation. To control for potential residual spatial trends and confounding, we included spatial smoothers within the model using natural cubic splines with 5 degrees freedom for both county centroid latitude and longitude. We further calculated Moran’s I of the standardized residuals of tri-pollutant main models for each state, to examine the presence of spatial autocorrelation in the residuals.

### Data sources on covariates

We adjusted for three county-level healthcare capacity covariates, including the number of intensive care unit (ICU) beds, hospital bed, and active medical doctor per 1000 people. Number of ICU beds were based on Kaiser Health News analysis of 2018 and 2019 hospital cost reports filed to the Centers for Medicare & Medicaid Services. Numbers of active medical doctors and hospital beds of 2017 were obtained from the Area Health Resources Files. State-level number of COVID-19 tests performed up to April 29, 2020 was derived from the Covid Tracking Project, based on which we calculated the positive rate in each state, i.e. the percentage of tests performed that are positive for COVID-19. The travel distance mobility data were released from the Descartes Labs and mapped by the GeoDS Lab using anonymized location data from smartphones (Warren and Skillman, 2020; Gao et al., 2020). The travel mobility index was a measure to compare the daily individual-level travel distance pattern to that in February. To enhance privacy, individual data are de-identified and aggregated to the county level. We calculated the county-level mean mobility index from March 1, 2020 to April 29, 2020 to represent the dramatic mean human mobility changes in reaction to the COVID-19. County-level socioeconomic status (SES) in 2015 was measured by social deprivation index, which is a composite measure of area-level deprivation based on seven characteristics, including income, education, employment, housing, household characteristics, transportation, and demographics. SDI has commonly served as an area-level composite measure of SES in other studies of health and health outcomes. County-level sociodemographic covariates in 2017 such as percentage of elderly (age≥60) and percentage of male were derived from Area Health Resource Files, and population density was derived from the 2018 US Census. County-level behavioral risk factors, including population mean BMI (an indicator of obesity) and percentage of ever smokers, were derived from the 2011 US CDC Behavioral Risk Factor Surveillance System (BRFSS). From Phase 2 of the North American Land Data Assimilation System (NLDAS-2), we acquired hourly 1/8th degree gridded near-surface air temperature and specific humidity data from January 22, 2020 through April 29, 2020 (Xia et al., 2012), based on which we calculated the mean temperature and relative humidity for each 1/8th degree grid. We linked each county’s centroid to the nearest 1/8th degree grid and assigned the mean temperature and relative humidity.

### Sensitivity analyses

We also conducted a series of sensitivity analyses to test the robustness of our results to outliers, confounding adjustment, and epidemic timing (Supplementary Appendix Figures S1 and S2). Given that New York city has far higher COVID-19 cases and deaths than any other regions in the US, which can be a very influential observation, we excluded all five counties within New York city and repeated the analysis. In another set of sensitivity analyses, we restricted the study only to the most recent 4 weeks (April 1 to April 29), when the case count and death count may be more reliable and accurate than earlier periods and when COVID-19 tests were more available. We also conducted sensitivity analysis by using air pollution data averaged between 2000 to 2016. To assess the impact of potential bias of individual covariates, we fit models by omitting a different set of covariates for each model iteration while comparing effect estimates. Statistical tests were 2-sidedand statistical significance was determined with an alpha of 0.05. All statistical analyses were conducted used R version 3.4.

## Notes

### Competing Interest Statement

The authors have declared no competing interest.

